# Comparative immunogenicity and effectiveness of mRNA-1273, BNT162b2 and Ad26.COV2.S COVID-19 vaccines

**DOI:** 10.1101/2021.07.18.21260732

**Authors:** Vivek Naranbhai, Wilfredo F. Garcia-Beltran, Christina C. Chang, Cristhian Berrios Mairena, Julia C. Thierauf, Grace Kirkpatrick, Maristela L. Onozato, Ju Cheng, Kerri J. St. Denis, Evan C. Lam, Clarety Kaseke, Rhoda Tano-Menka, Diane Yang, Maia Pavlovic, Wendy Yang, Alexander Kui, Tyler E. Miller, Michael G. Astudillo, Jennifer E. Cahill, Anand S. Dighe, David J. Gregory, Mark C. Poznansky, Gaurav D. Gaiha, Alejandro B. Balazs, A. John Iafrate

## Abstract

**Background:** Understanding immunogenicity and effectiveness of SARS-CoV-2 vaccines is critical to guide rational use.

**Methods:** We compared the immunogenicity of mRNA-1273, BNT-162b2 or Ad26.COV2.S in ambulatory adults in Massachusetts, USA. To correlate immunogenicity with effectiveness of the three vaccines, we performed an inverse-variance meta-analysis of population level effectiveness from public health reports in >40 million individuals.

**Results:** A single dose of either mRNA vaccine yielded comparable antibody and neutralization titers to convalescent individuals. Ad26.COV2.S yielded lower antibody concentrations and frequently negative neutralization titers. Bulk and cytotoxic T-cell responses were higher in mRNA1273 and BNT162b2 than Ad26.COV2.S recipients, and <50% of vaccinees demonstrate CD8+ T-cell responses to spike peptides. Antibody concentrations and neutralization titers increased comparably after the first dose of either vaccine, and further in recipients of a second dose. Prior infection was associated with high antibody concentrations and neutralization even after a single dose and regardless of vaccine. Neutralization of beta, gamma and delta strains were poorer regardless of vaccine. Relative to mRNA1273, the effectiveness of BNT162b2 was lower against infection and hospitalization; and Ad26COV2.S was lower against infection, hospitalization and death.

**Conclusions:** Variation in the immunogenicity correlates with variable effectiveness of the three FDA EUA vaccines deployed in the USA.

## Introduction

Prophylactic vaccines against SARS-CoV-2 are being deployed globally to combat the COVID-19 pandemic. A burgeoning body of evidence links the immunogenicity of the different vaccines to the degree of protection from infection or disease, although a precise correlate has not been agreed upon and studies of direct comparisons of vaccines are limited. Vaccination with mRNA-1273[1] (100ug, Moderna), BNT162b2[2] (30ug, Pfizer) and Ad26.COV2.S[3] (5×10^10^ viral particles, Johnson & Johnson/Janssen) were each shown to be efficacious in reducing the risk of severe disease and infection in randomized clinical trials[1–3], and have received Emergency Use Authorization (EUA) or full approval from the United States FDA. All three vaccines encode a largely similar SARS CoV-2 spike protein homologous to the SARS-CoV-2 strain isolated in Wuhan (China) but differ in dose and mechanism of delivery (mRNA vs. adenovirus vectored). Scale-up of vaccination in the general population may plausibly result in outcomes different to the original trials because of broader inclusion in the real world, the evolution of viral variants and potential variation in vaccine production. Immunogenicity analyses nested within the randomized trials of several vaccines have consistently demonstrated a quantitative association between anti-spike antibodies and/or neutralization titers and infection outcomes[4–7]. In animal models, experimental transfer of antibodies protects from infection[8–10] Click or tap here to enter text. and in human randomized controlled-trials, prophylactic administration of neutralizing antibodies reduces incidence of clinical COVID-19[11,12]. However, there is no consensus between vaccine manufacturers on the immune assays to employ, nor in the details of similar assays, for example which viral variant to assess neutralization against. There are few data regarding the comparative immunogenicity of these vaccines, except for indirect inferences from publicly available trial data from manufacturers[13–15] and small recent studies[13]. These factors collectively justify direct comparisons between vaccines in real world settings using consistent methods. Such studies may provide data to base decisions regarding which vaccine to deploy, and timing of additional ‘booster’ doses.

We compared the immunogenicity of mRNA1273, BNT162b2 and Ad26.COV2.S during the first months following their deployment in the pandemic in the USA. We found a distinct hierarchy in humoral and cellular immunity, including towards currently circulating viral variants. Moreover, we find that this hierarchy is mirrored by variation in population scale effectiveness.

## Methods

### Cohort description

Use of human samples was approved by Partners Institutional Review Board (protocol 2020P001081 and 2020P002274). Consenting adults in Chelsea, Massachusetts were enrolled in a study of antibody responses and sampled in August 2020 and/or early 2021 (March or April 2021). Data in this study are also derived from previously reported cohorts of healthy adults who had received vaccination and enrolled in a COVID vaccine biobanking study. Demographic data, information regarding prior SARS CoV-2 testing, symptoms, and exposure was collected as was vaccine related information. Pre-pandemic serum samples were obtained from the clinical laboratories at Massachusetts General Hospital (MGH) as previously described[17]. We grouped patients by time post vaccination using a 7 day window to capture the expected kinetics of antibody production after vaccination. Individuals who had received a single dose of vaccination >=7 days prior (or had received a second dose less than 7 days prior) were analyzed as one-dose recipients; those who had received two doses >=7 days prior were analyzed as two-dose recipients.

### Antibody assays

Individuals were defined as having evidence of prior SARS CoV-2 infection if they had either a prior positive nucleic acid test or a measurable anti-nucleocapsid antibody on the Roche Elecsys assay run at the Massachusetts General Hospital Core clinical lab, a Clinical Laboratory Improvement Amendments (CLIA) laboratory.

Total anti-spike (IgA/M/G) antibody was measured on the Roche Elecsys Anti-SARS-CoV-2 immunoassay according to the manufacturer’s instructions. An antibody binding cut-off index (COI) >0.8 was defined as positive per the manufacturer’s guidance. The assay is semi-quantitative and values >2,500 U/ml triggered additional dilution (where sample availability allowed) to yield titers up to 250,000 U/ml. All assays were run blinded to clinical information such as vaccine type and dose.

Quantitative detection of total (IgA/M/G) and individual isotype (IgG, IgA or IgM) antibodies to SARS-CoV-2 receptor binding domain (RBD) was performed by enzyme-linked immunosorbent assay (ELISA) as previously described[17] with the only modification being adaptation of the protocol to 384-well plates.

Neutralization was measured using a SARS-CoV-2 pseudovirus neutralization assay that has been previously described[14]. Briefly, lentiviral particles encoding both luciferase and ZsGreen reporter genes were pseudotyped with SARS Cov-2 spike protein and produced in 293T cells, titered using ZsGreen expression by flow cytometry and used in an automated neutralization assay with 50–250 infectious units of pseudovirus co-incubated with three-fold serial dilutions of serum for 1 h. Neutralization was determined on 293T-ACE2 cells. Percent neutralization was determined by subtracting background luminescence measured in cell control wells (cells only) from sample wells and dividing by virus control wells (virus and cells only). Data was analyzed using GraphPad Prism and pNT50 values were calculated by taking the inverse of the 50% inhibitory concentration.

ELISAs against multimeric RBDs from SARS CoV-2 (αα319-591), OC43 (αα315-675) and HKU1 (αα307-675) were prepared using 8×His tagged protein expressed in HEK 293 cells. Plasma was diluted 1:1000 (HKU1) or 1:2000 (SARS-CoV-2 and OC43) and detected using HRP-linked, goat anti-Human Fc gamma (Cell Signaling Technology).

### ELISPOT assays

Interferon gamma (IFN-γ) ELISpot assays were performed as previously described[15] according to the manufacturer’s instructions (Mabtech). 500,000 PBMCs per test were incubated with SARS-CoV-2 peptide pools at a final concentration of 1□μg/□ml for 16– 18h. CEF peptide pool (Mabtech; 1ug/mL), anti-CD3 (Clone OKT3, Biolegend, 1ug/mL) and anti-CD28 Ab (Clone CD28.2, Biolegend, 1ug/mL) were used as positive controls. To quantify antigen-specific responses, mean spots of the DMSO control wells were subtracted from the positive wells, and the results were expressed as spot-forming units (SFU) per 10^6^ PBMCs. Responses were considered positive if the results were >10□SFU/10^6^ PBMCs following control subtraction.

### Meta-analysis of effectiveness data

We systematically searched pubmed, and recent online news (including twitter) for mention of reports of breakthrough infection according to vaccine type using the terms ‘breakthrough’, ‘effectiveness’, ‘Moderna’, ‘mRNA1273’, ‘Pfizer’, ‘BNT162b2’, ‘J&J’ or ‘Ad26.COV2.S’. Studies reporting breakthrough case rates, hospitalization or mortality for a clearly defined (denominator) population, stratified by vaccine type for all three vaccines, were eligible for inclusion. We identified five eligible sources of breakthrough infection rates. In some instances, the absolute number of breakthrough cases was reported, and the denominator (number of recipients of each particular vaccines) was abstracted from a secondary source. We detail these in the online materials. None provided individual level data to adjust for covariates. Inverse-variance meta-analysis was performed in R using the metagen function.

### General statistical methods

We performed multivariate linear regression in R (v4.05) using the lm function with log_10_ transformed spike values or pNT50 as the dependent variable, and age, sex, days post vaccination, or vaccine group as the independent variables. Graphics were rendered in Prism v9.0. We modelled kinetics of vaccine response by plotting the individual measures or repeated measures (for donors who had repeated measures performed), and calculating a trend line using the R geom_smooth() function with span 0.1.

## Results

We characterized the immunogenicity of mRNA-1273 (Moderna), BNT162b2 (Pfizer-BioNTech) and Ad26.COV2.S (Johnson & Johnson (Janssen)) in ambulatory adults enrolled in a community study of healthy individuals in Chelsea, Massachusetts and a biobanking effort among laboratory or health care workers in Boston, Massachusetts. In total we include data from 215 participants who had received one (*n* = 99) or two doses (*n* = 116) of vaccine ≥7 days prior, 130 unvaccinated participants with asymptomatic or symptomatic prior infection confirmed by positive anti-nucleocapsid antibody, 112 uninfected (anti-nucleocapsid antibody negative), and 1,220 historical controls sampled before the pandemic[16]. The median age of vaccinated participants was 39 years (IQR 31-55 years) and 120/215 (56%) were female. We compared immune responses according to prior infection, and vaccination type and dose and adjusted for age, sex and duration after vaccination in all analyses.

### Binding antibody response to vaccination

We assessed total IgG/M/A binding antibody levels against the SARS-CoV-2 spike protein (Roche Elecsys Anti-SARS-CoV-2 S), and found substantial variation in antibody concentrations depending on prior infection, vaccine type and vaccine dose (**Figure 1A**, summarized in a multivariate regression model in **Supplementary Table 2)**. Among participants without prior infection, antibody concentrations after a single dose of mRNA-1273 was comparable to convalescent individuals (geometric mean concentration in U/mL (GMC) 222 vs. 189, adjusted *p*=0.4 for mRNA-1273) and after a single dose of BNT162b2 was lower than convalescent individuals (GMC 71 vs. 189, adjusted *p*=0.01 for BNT162b2). Recipients of Ad26.COV2.S without prior infection had an approximately 25-fold lower antibody concentrations than convalescent unvaccinated individuals (GMC 6.9 vs 189, adjusted *p* <0.001). Participants with no prior infection who received a single dose of either of the three vaccines were sampled at a comparable duration after vaccination (a median of 22-24 days). A single dose of mRNA-1273 or BNT162b2 induced higher titers than Ad26.COV2.S (adjusted *p* < 0.001 for both comparisons, **Supplementary Table 3**). At a median of 24 days post-vaccination, 27.3% (6/22) of Ad26.COV2.S recipients had undetectable antibody levels. Notably, none of 22 Ad26.COV2.S recipients had a major medical comorbidity or received immunosuppressive medications in the prior six months. Receipt of both doses of mRNA-1273 or BNT162b2 were associated with substantially higher antibody concentrations than convalescent individuals (GMC 6486 for mRNA-1273 and GMC 2455 for BNT162b2 vs. 189, adjusted *p*<0.001 for both comparisons). Individuals with prior infection who were vaccinated had ∼2 log_10_ IU/ml higher antibody concentrations than convalescent individuals regardless of vaccine type, even after a single dose.

**Figure 1:**
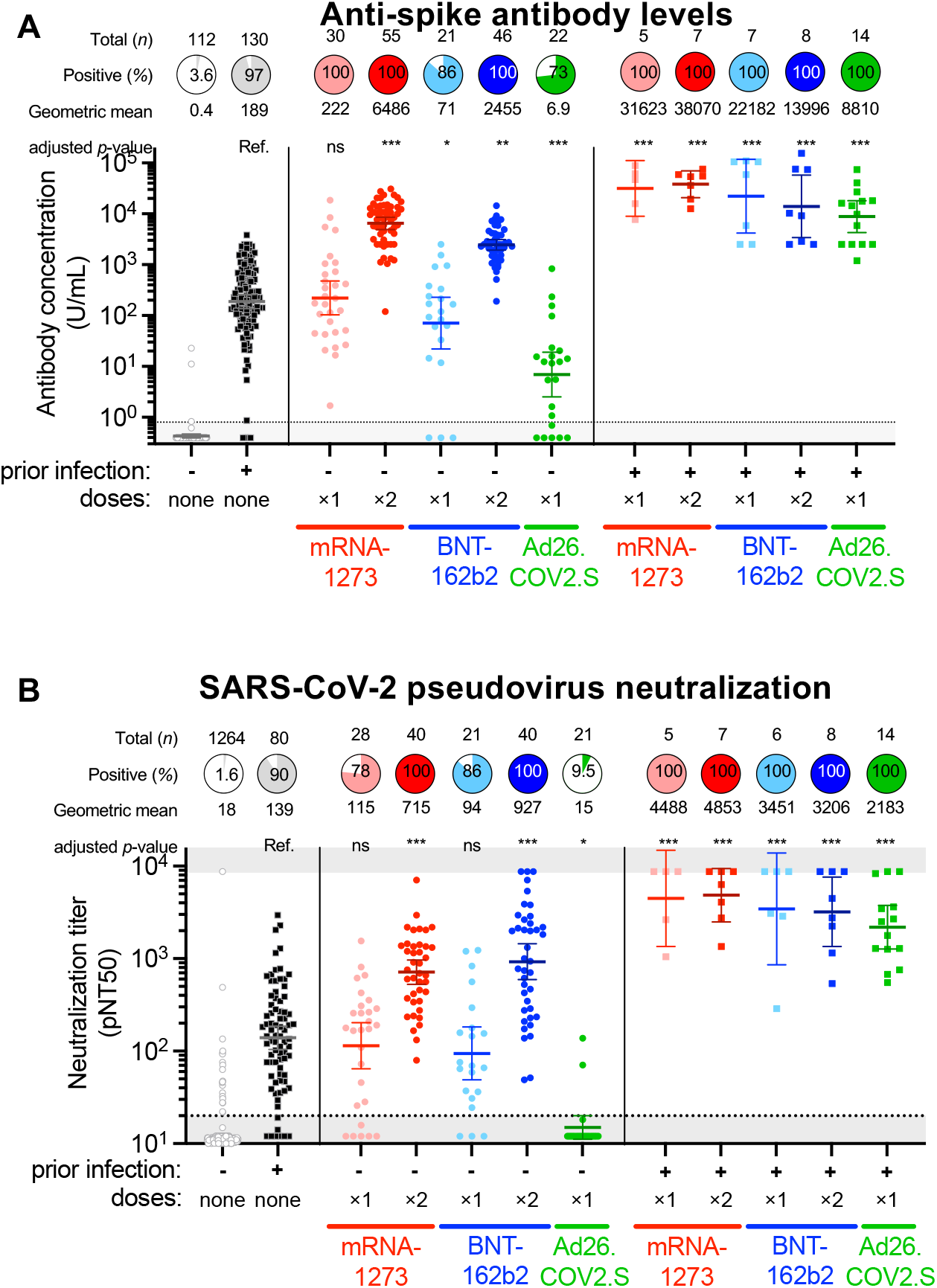
Immunogenicity of mRNA-1273, BNT-162b2 and Ad26.COV2.S in individuals with and without prior SARS-CoV-2 infection. **Panel A** shows the quantitative SARS-CoV-2 spike IgG/A/M antibody concentration (Roche Elecsys Anti-SARS-CoV-2 assay) in U/ml of serum for 112 participants without prior infection or vaccination, 130 with prior infection and 215 following vaccination. An antibody titre of 0.8 U/ml was considered positive (dotted line). The number of donors, proportion positive and geometric mean concentration is shown above each group. **Panel B** shows pseudovirus neutralization titer 50 (pNT50, defined as the titer at which the serum achieves 50% neutralization of SARS-CoV-2 wildtype pseudovirus entry into ACE2 expressing 293T cells) for a subset of the donors above and an additional 1220 pre-pandemic controls (from Garcia-Beltran et al.[18]) used in assay validation and deriving the cutoff shown. The threshold for defining positive individuals, denoted by the dotted horizontal line, was a pNT50 of 20[18]. The number of donors, proportion positive (at a threshold of 1:20) and geometric mean titer is shown above each group. In both panel A and B, for each group the horizontal line denotes the geometric mean concentration, and whiskers extend to 95% confidence interval. Asterisks denote p-values adjusted for age, sex and duration after vaccination at time of sampling, relative to unvaccinated individuals with prior infection (see supplementary Table 2 and 4) as follows: ns not significant, * *p*<0.05, ** *p*<0.01, *** *p*<0.001. :

We next sought to confirm these results using orthogonal assays. In an independent, validated total IgG/M/A or IgG ELISA, measurement of receptor binding domain (RBD) binding antibodies confirmed the differences above; and IgM and IgA responses were low (**Figure S1**). Further measurement of IgG against RBD-multimers also confirmed these findings (**Figure S2**). Control experiments revealed that IgG responses against equivalent RBD-multimers of less pathogenic, common coronaviruses OCU43 and HKU1 were comparable between groups supporting the specificity of these findings, and controlling for any sample collection or storage artifacts between recipients of the three vaccines (**Figure S3**).

### Neutralizing antibody responses to vaccination

We assessed the ability of serum from participants to neutralize SARS-CoV-2 in using a well-characterized assay[14,16] that utilizes lentivirus pseudotyped with the Spike protein of the Wuhan isolate of SARS-CoV-2. Neutralization data was available for 35 Ad26.COV2.S vaccinees and a 80 mRNA-1273 and 75 BNT162b2 vaccinees. Among pre-pandemic and confirmed uninfected individuals, a neutralization titer (pNT50) cutoff of 20 identified 1.6% of samples as positive. Using this threshold, 90.1% of unvaccinated convalescent individuals demonstrated neutralization (**Figure 1B**). Following a single dose of mRNA1273 or BNT162b2, neutralization titers were comparable to convalescent individuals (geometric mean titer (GMT) 115 for mRNA1273 and 94 for BNT162b2 vs. 139 for convalescent donors, *p*=ns for both comparisons **Supplementary Table 4**) and 78% and 86% neutralized virus. Titers were higher after both doses of mRNA vaccine (GMT 715 for mRNA1273 and 927 for BNT162b2, adjusted *p*<0.001 for both), and serum from all two-dose mRNA vaccinees neutralized wild-type SARS-CoV-2 pseudovirus. In contrast, 9.5% (2/21) Ad26.COV2.S recipients had a neutralization titer >20. Among individuals with prior infection, receipt of one or two doses of either of the three vaccines generated high neutralization titers that ranged from 46-times higher (for one dose Ad26.COV2.S) to 200-times higher (for two dose mRNA1273) than unvaccinated individuals with prior infection.

### T-cell responses to vaccination

To explore differences in cellular immune response to vaccination, we assessed T-cell responses to SARS-CoV-2 spike peptides (wild-type strain) by IFN-γ ELISpot in 29 unvaccinated individuals without and 26 with prior infection, and 36 vaccinees without prior SARS CoV-2 infection who completed their vaccination schedule (**Figure 2**). We first examined bulk T-cell responses (which include both CD4+ and CD8+ T-cells) (**Figure 2A**). For reference, 92% of unvaccinated participants with prior infection had a measurable response with a mean of 164 SFU per 10^6^ PBMC. Spike-specific bulk T-cell responses varied significantly by vaccine (Kruskall-Wallis p=0.007). Bulk T-cell responses were higher, and more frequent in recipients of mRNA-1273 or BNT-162b2 than Ad26.COV2.S (mean 511 or 348 vs. 97 SFU/10^6^ PBMC, *p*=0.003). Relative to convalescent individuals (adjusting for age and sex) the magnitude of response was higher in mRNA1273 recipients (effect estimate 310 SFU/10^6^ PBMC, 95% CI 90-530; p=0.007), tended towards being significantly higher in BNT162b2 recipients effect estimate 182 SFU/10^6^ PBMC, 95% CI 28-391; p=0.08), and was non-significantly lower after Ad26.COV2.S (effect estimate -62 SFU/10^6^ PBMC, 95% CI -239-114; p=0.05).

**Figure 2:**
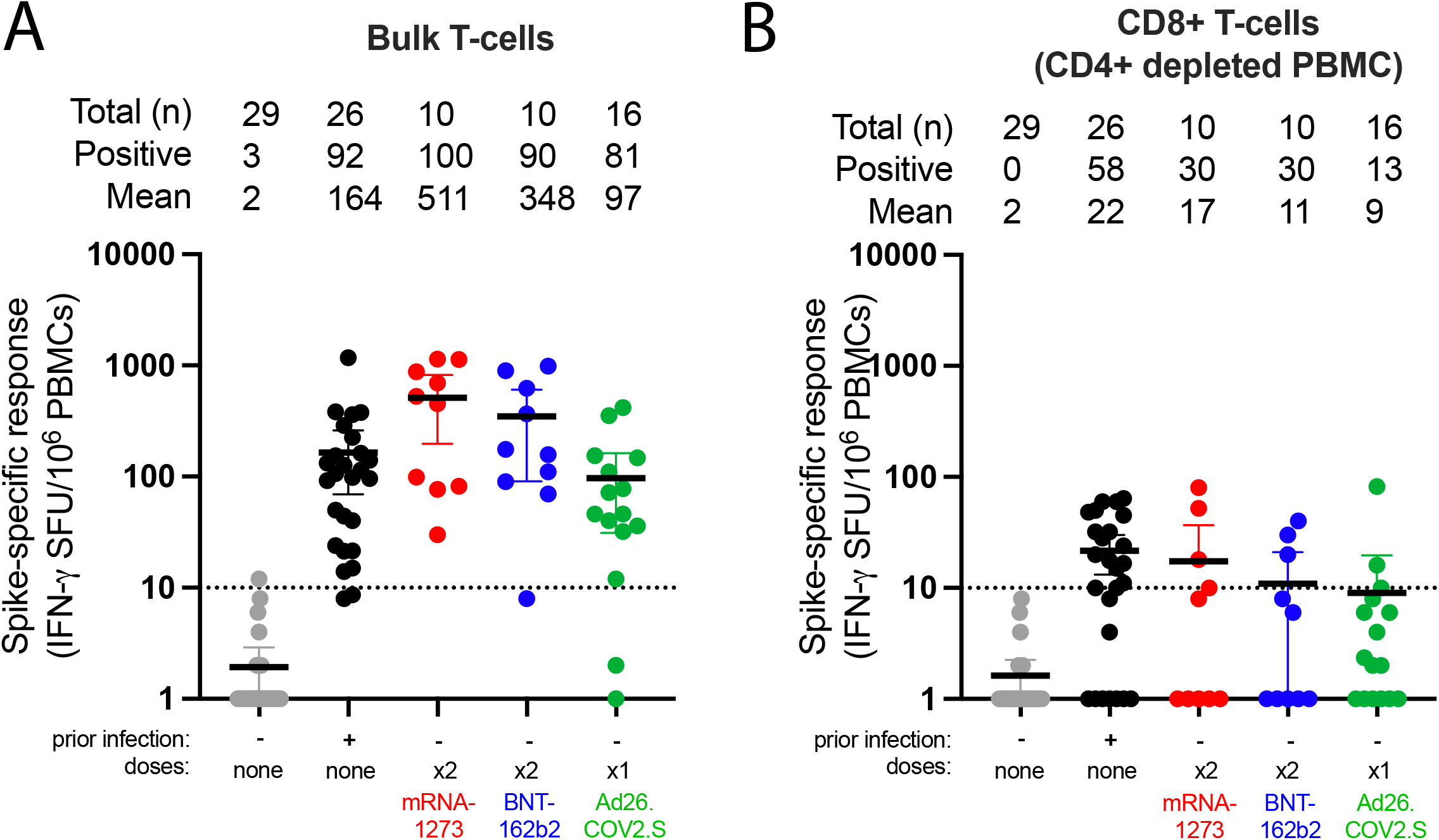
**T-cells** responses to spike peptides after. **mRNA-1273, BNT-162b2 and Ad26.COV2.S in individuals without prior SARS-CoV-2 infection measured in bulk T-cells (Panel A), and CD4-depleted T-cells (Panel B)**. The threshold for defining positive individuals, is denoted by the dotted horizontal line at 10 SFU/10^6^ PBMC. Data are normalized to background DMSO control responses. Asterisks denote p-values adjusted for age, sex and duration after vaccination at time of sampling, relative to unvaccinated individuals with prior infection (see supplementary Table 2 and 4) as follows: ns not significant, * *p*<0.05, ** *p*<0.01, *** *p*<0.001.

To evaluate anti-spike cytotoxic T-cell responses we measured spike-specific responses after depleting CD4+ cells (**Figure 2B**). Unvaccinated participants with prior infection had a mean 22 SFU/10^6^ CD4+ depleted PBMC, and 58% had a measurable response. Regardless of vaccine, fewer participants had measurable cytotoxic T-cell responses to spike peptides (30% for mRNA1273, 30% for BNT162b2 and 13% for Ad26.COV2.S) relative to convalescent individuals (58%). The magnitude of responses were higher among recipients of mRNA-1273 (or BNT-162b2) than Ad26.COV2.S (mean 17 or 11 vs. 9 SFU/10^6^ PBMC, adjusted *p*=0.04). Neither bulk nor CD4-depleted spike-specific responses correlated with binding antibody titers (data not shown).

### Change in antibody responses over time

To explore how antibody titers vary over time, we performed two additional analyses. First, to assess whether an increase in anti-SARS-CoV-2 antibodies occurs at later timepoints, which has been described among Ad26.COV2.S recipients[3,17], we obtained repeat measures in a subset of 15 Ad26.COV2.S recipients without prior infection. For this subset the baseline sampling was at a median 23 (range 7-44) days and the follow-up sampling at a median 66 (range 25-82) days after vaccination. Anti-spike antibody concentrations increased among all Ad26.COV2.S individuals (**Figure 3A**); however, only four individuals had an increase in neutralization, and 73% (11/15) remained with a pNT50 <20 (**Figure 3B**). Second, we pooled initial and repeat measures among individuals without prior infection and modelled the kinetics of responses for all three vaccines. Over the first ∼6 weeks following receipt of first dose of vaccination, antibody concentrations increased regardless of vaccine (**Figure 3C**) but the estimated plateau titers were lower among Ad26.COV2.S recipients. Since Ad26.COV2.S is administered as a single dose, we decided for fair comparison to analyze the kinetics of response for the three vaccines in the period just after the first doses and prior to the second mRNA vaccines (i.e. the first four weeks for BNT162b2 and five weeks for mRNA1273). Titers increased for all three vaccines, and the rate of increase was not statistically dissimilar between vaccines (interaction *p*=0.85).

**Figure 3:**
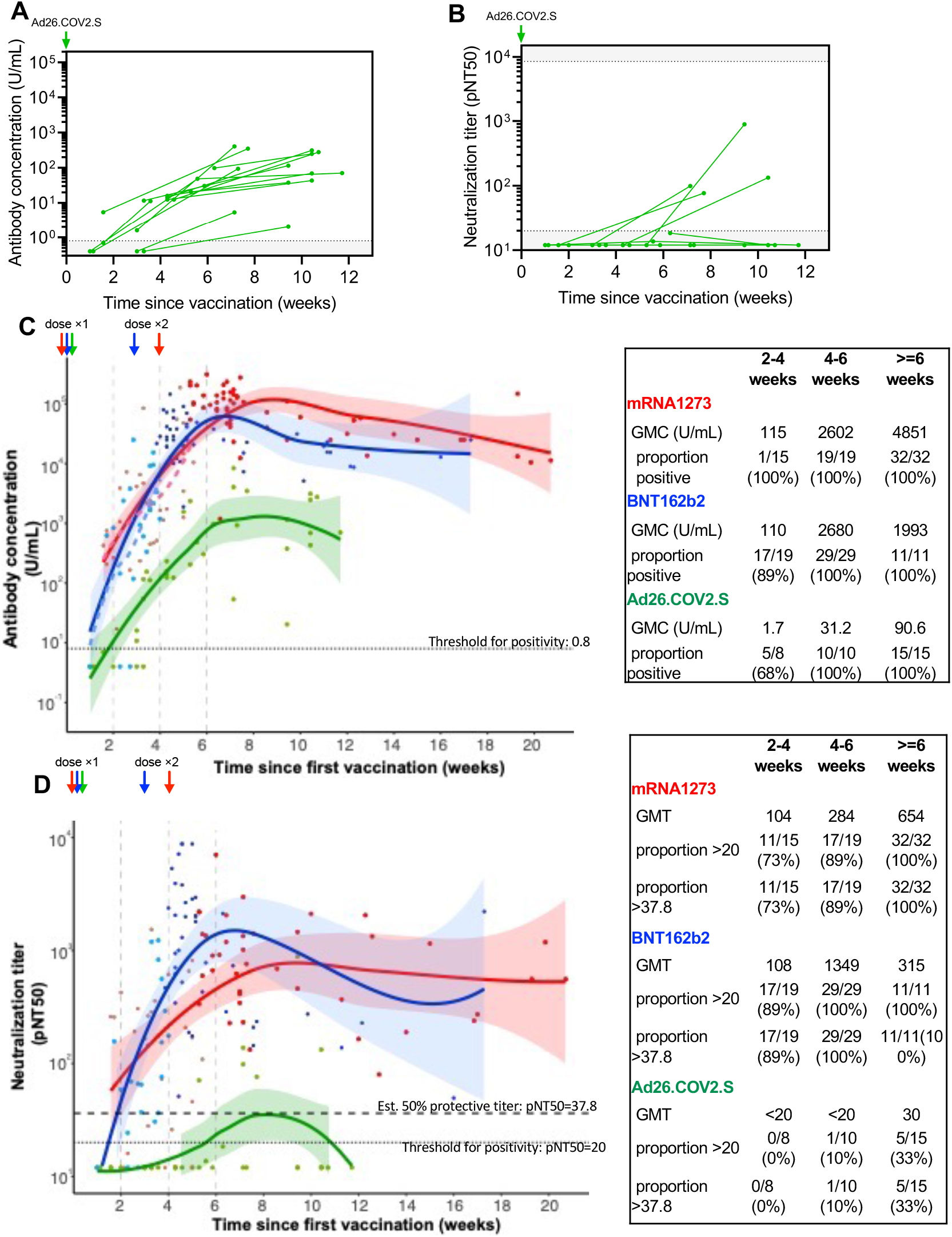
Follow-up measurement of response in Ad26.COV2.S recipients and kinetics of humoral responses to mRNA1273, BNT162b2 and Ad26.COV2.S SARS-CoV-2 vaccines. Longitudinal assessment of SARS-CoV-2 spike IgG/A/M antibody titers (**Panel A)** and virus neutralization **(Panel B)** in 15 Ad26CoV2.S vaccinees with baseline and repeat measures. Pooling all data in, we modelled the kinetics of antibody concentration and neutralization according to vaccine. **Panel C** shows SARS-CoV-2 spike IgG/A/M antibody levels and **Panel D** shows virus neutralization for all donors sampled after vaccination, with best-fit lines (Loess fit) shown and colored according to vaccine type, over the first 20 weeks following vaccination. In Panels C and D, the geometric mean concentration or titer, and proportion positive at threshold indicated are shown for the periods 0-2, 2-4, 4-6 and >=6 weeks in the data table insert to the right of the main figure. Each individual point and corresponding fit are colored according to vaccine type (red mRNA-1273, blue BNT162b2 or green Ad26COV2.S), the shaded area adjacent to each line denotes the 95% confidence interval, and points are additionally shaded according to dose. Dashed lines show best-fit lines for the period after receipt of the first dose of mRNA-1273 or BNT162b2 in Panel C. In Panel D, the dotted line denotes a pNT50 threshold of 20 derived from study of pre-pandemic controls, and the upper dashed line denotes a pNT50 titer of 37.8 which represents 20% of the geometric mean neutralization titers of unvaccinated individuals with prior infection and corresponds with 50% estimated protection in Khoury et al[4].

Analysis of the kinetics of viral neutralization show similar shapes and time-course as the antibody assay for mRNA-1273 and BNT162b2, with peak values reached 6-10 weeks post first vaccine (**Figure 3D**). Such a comparison was not possible for Ad26.COV2.S recipients since most did not neutralize virus at any timepoint. Prior publications have estimated that 20% of the GMT value of convalescent individuals is a good threshold for predicting titers offering ∼50% protection from reinfection[4]. In this cohort, 20% of the GMT of convalescent individuals is 27.8, and we used this threshold to show that most recipients of mRNA-1273 (73%, 11/15) or BNT162b2 (89%, 17/19) achieved predicted protective neutralization titers rapidly, before receipt of the second dose of vaccination, and show that these are sustained for several months.

Most reported neutralization data amongst vaccinated individuals are derived from vaccine manufacturer’s study of trial participants. The assays used, in particular the virus strain, varies between studies. Notably studies of Ad26.COV2.S recipients utilized the Victoria strain of SARS CoV-2 [14], a strain that has been noted to be more easily neutralized[18]. We introduced the Victoria strain-associated S247R mutation into the SARS CoV-2 Wuhan isolate used in this study. Sera from vaccine recipients demonstrated higher neutralization titers against this S247R-containing variant than the original Wuhan strain regardless of vaccine administered (**Figure S4**). Therefore differences in the described neutralization seen in this study vs. the Ad26.COV2.S trialsClick or tap here to enter text. may be partially accounted for by use of the S247R viral variant in prior studies.

### Neutralization of viral variants

Variants of SARS CoV-2 have arisen over the course of the pandemic. mRNA1273, BNT162b2 and Ad26.COV2.S each encode the wild-type (Wuhan strain) Spike protein as the sole viral antigen. We evaluated vaccine-mediated neutralization of the beta (B.1.351), gamma (P1) and delta (B.1.617.2) variants of concern and SARS-COV-1 by serum from volunteers who completed a vaccine series. The overall neutralization of each variant was similar between both mRNA vaccines in that neutralization of the beta variant was markedly impaired relative to the ancestral Wuhan strain (30-fold reduced GMT for mRNA1273 and 48-fold reduced GMT for BNT162b2), while neutralization of both gamma and delta variants was modestly impaired (**Figure 4**). Serum from Ad26.COV2.S recipients demonstrated low neutralization for all strains, as most individuals had neutralization that was not measurable.

**Figure 4:**
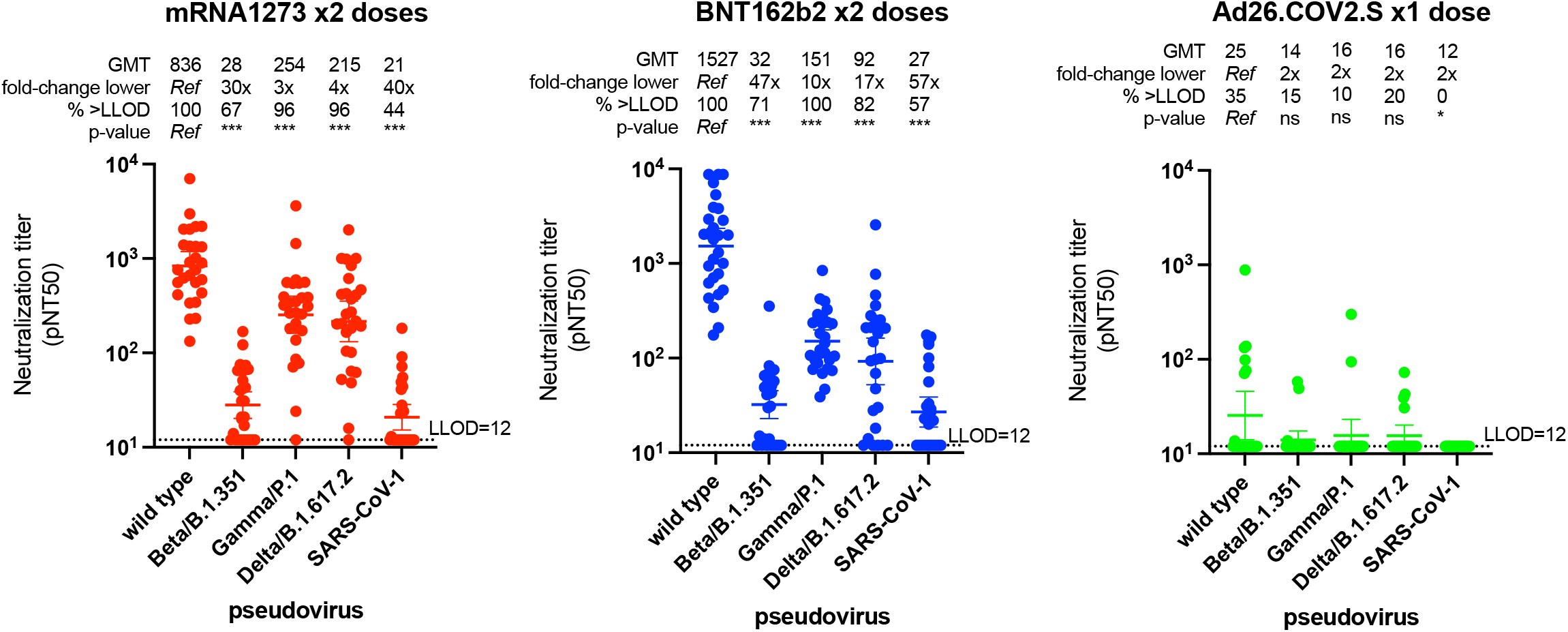
Neutralization of SARS CoV-2 variants of concern by sera from healthy donors without prior infection vaccinated with mRNA1273 x2 dose (**Panel A, n=27**), BNT162b2 x2 dose(**Panel B, n=28**) or Ad26.COV2.S x1 dose (**Panel C, n=20**). The pseudovirus neutralization titer 50 (pNT50, defined as the titer at which the serum achieves 50% neutralization of the relevant SARS-CoV-2 wildtype pseudovirus entry into ACE2 expressing 293T cells) is shown for each donor against the SARS CoV-2 wild type (Wuhan) strain, the beta gamma and delta viral variants and SARS CoV-2. The geometric mean titer, fold-change relative to neutralization of wild-type, and proportion above the lower limit of detection (LLOD denoted by horizontal dotted line at pNT50=12), and degree of statistical evidence relative to wild-type is shown above each group. Non-parametric (Friedman) test p-values relative to wild-type are denoted as ns not significant, * *p*<0.05, ** *p*<0.01, *** *p*<0.001. For each group the horizontal line denotes the geometric mean titer, and whiskers extend to 95% confidence interval.

### Comparison of population-level vaccine effectiveness mirrors differences in immunogenicity

We hypothesized that the observed relative differences in immunogenicity between the three studied vaccines may correlate with their relative clinical effectiveness against infection and severe disease (hospitalization and/or death) at the population-level. In randomized-clinical trials, performed prior to heterologous variants rising to dominate the pandemic, and conducted largely among younger adults without major comorbidities, the primary end-point efficacy was 94.1% for mRNA1273[1], 94.6% for BNT162b2[2] and 66.1% for Ad26.COV2.S[3]. However, the differences between these studies with regard to study participant characteristics, study endpoints (severity of disease and onset relative to vaccination), the prevalence of viral variants and other factors make direct comparisons between vaccines difficult. We systematically searched for reports of comparative effectiveness, and performed a fixed-effects meta-analysis (**Figure 5**). A unique strength of examining population-level effectiveness of vaccines (as opposed to trial efficacy) is the ability to perform comparisons between vaccines. To mitigate concerns regarding selection and ascertainment biases we included only studies that report breakthrough infection, hospitalization and/or mortality stratified by vaccine for the entire population of a defined geographic region, reasoning that virus exposure, testing and reporting methods are typically consistent within a defined population and the denominator can be definitively estimated. We identified five sub-national (US states of California, Oklahoma and the District of Columbia) or national (Iceland and South Korea) sources of relative effectiveness data reporting: in summary, 102,604 breakthrough infections among 40,616,188 individuals who had completed the primary vaccine series (10,607,139 mRNA1273, 27,152,066 BNT162b2 and 2,856,983 Ad26.COV2.S). The reporting period largely covers recent months when the pandemic was dominated by the delta variant, but we note that the reports did not stratify rates by strain. Three sources reported 2,695 hospitalization and 298 deaths among 23,937,009 individuals (9,586,575 mRNA1273, 12,676,378 BNT162b2 and 1,674,056 Ad26.COV2.S). We note that the real-world effectiveness of each vaccine (relative to unvaccinated individuals) has been reported elsewhere and is high and consistent with the trial estimates, justifying global deployment of these vaccines and public health recommendations for universal vaccination and the goal of this analysis was a comparison between the vaccines.

**Figure 5:**
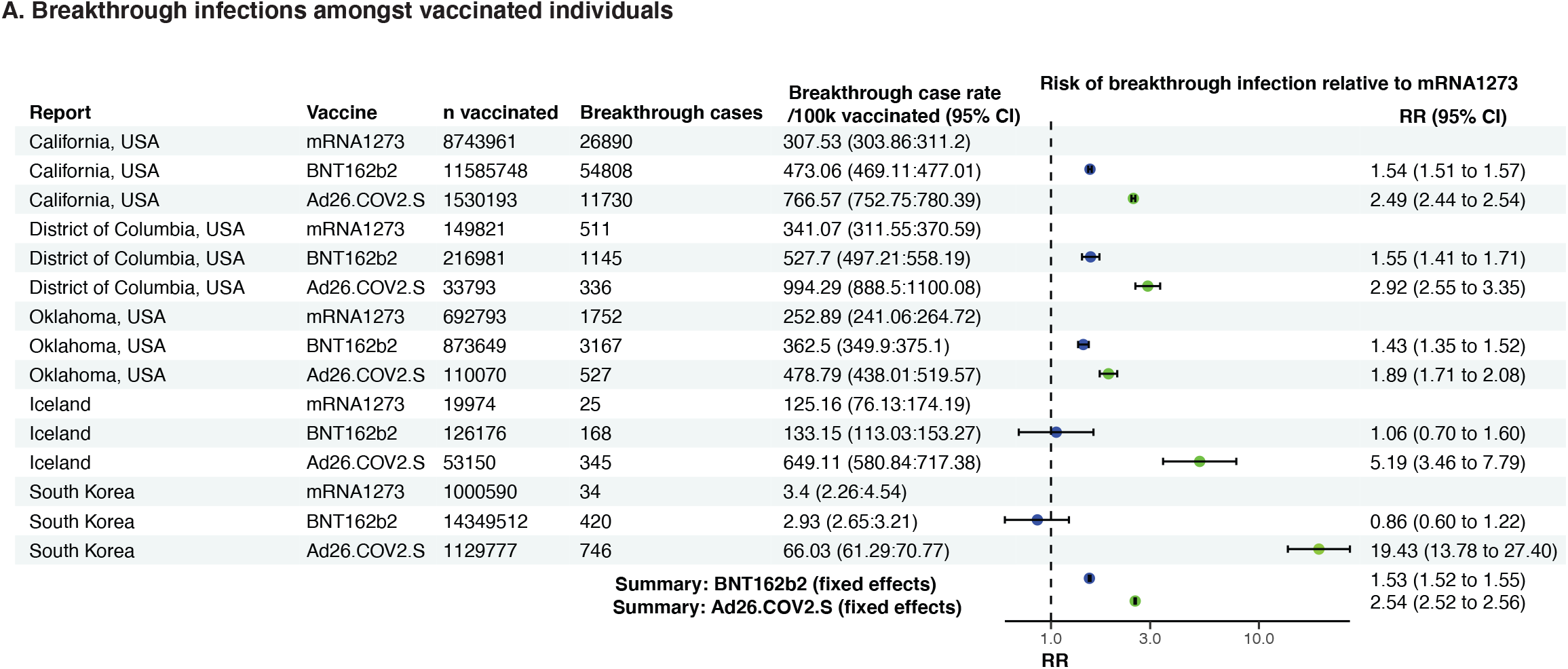
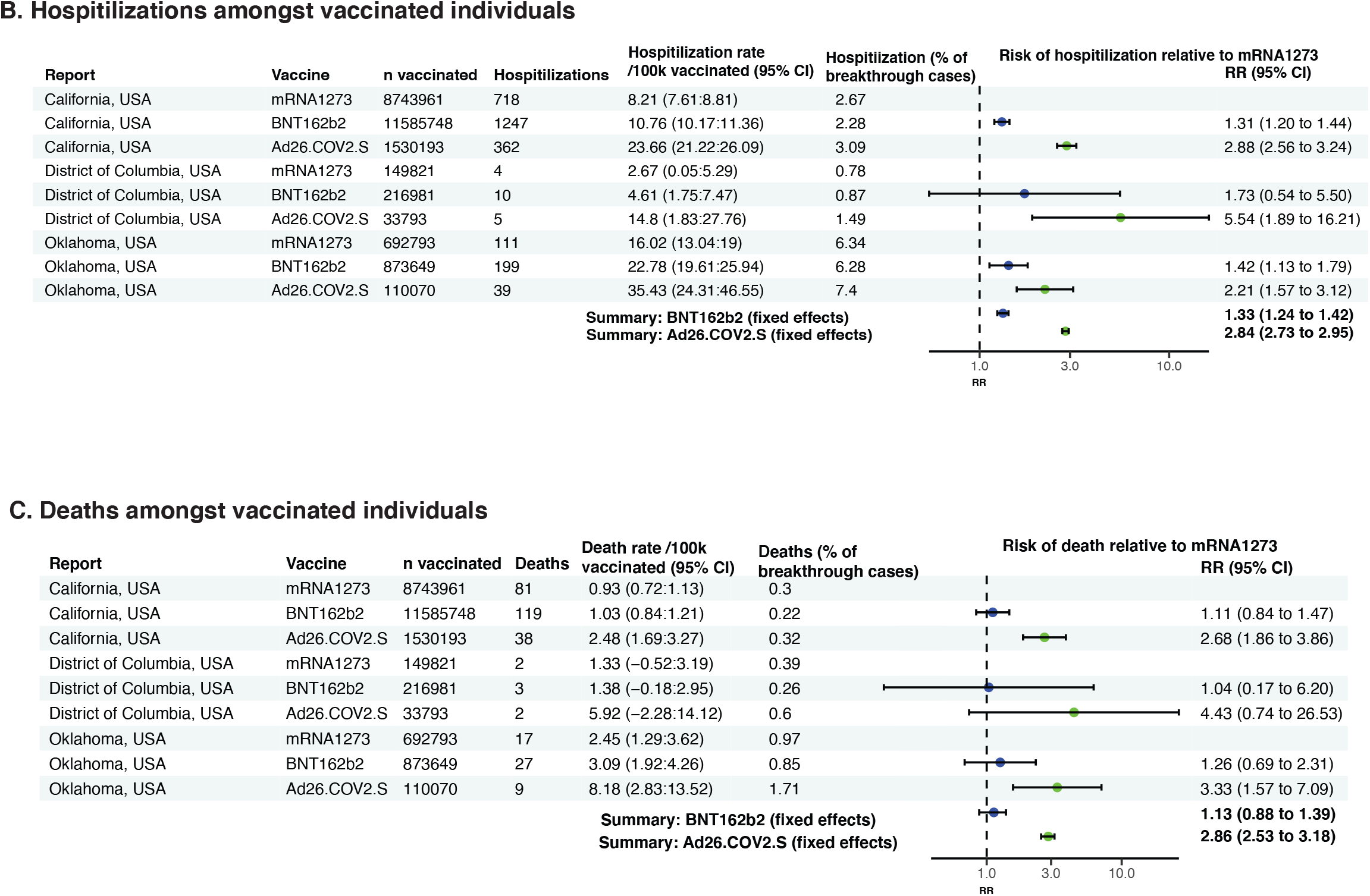
Forest plot of estimates from national or sub-national public health reports of comparative vaccine effectiveness against infection (Panel A), hospitalization (Panel B) or death (Panel C) for mRNA1273, BNT162b2 and Ad26.COV2.S recipients. See online materials for data sources.

In each of the five studies, rates of breakthrough infection were lowest after mRNA1273, higher after BNT162b2 and highest after Ad26.COV2.S, regardless of report although the absolute rate of breakthrough infection varied widely amongst the five reports. In a fixed-effects meta-analysis, the risk of breakthrough infection after BNT162b2 relative to mRNA1272 was 1.53 (95% CI 1.52-1.55), and after AD26.COV2.S was 2.54 (95% CI 2.52-2.56. The risk of hospitalization as a proportion of breakthrough cases were not markedly different between the three vaccines, but the absolute risk of hospitalization after BNT162b2 relative to mRNA1272 was 1.33 (95% CI 1.24-1.42), and after AD26.COV2.S was 2.84 (95% CI 2.73-2.95). Absolute death rates were low amongst vaccinated individuals consistent with very high levels of protection against death, regardless of vaccine. Overall mortality estimates were comparable between BNT162b2 and mRNA1273 recipients (RR 1.13 95% CI 0.88-1.39) but significantly higher after Ad26.COV2.S (RR 2.86 (2.53-3.18).

## Discussion

We studied immunogenicity in a healthy population of individuals receiving one of the FDA EUA vaccine during population scale-up. Regardless of immune measure examined namely anti-spike binding antibodies, anti-RBD binding antibodies, neutralization of wildtype, beta, gamma or delta strains, bulk T-cells or cytotoxic T-cells, we observed a consistent pattern with mRNA1273 being the most immunogenic followed by BNT162b2 and both mRNA vaccines being significantly more immunogenic than Ad26.COV2.S. Similarly, the effectiveness against infection, hospitalization or death in meta-analysis of population data was highest with mRNA1273, intermediate after BNT162b2 and lowest after Ad26.COV2.S. Differences in the efficacy[1–3] and effectiveness during deployment under emergency-use-authorization of mRNA-1273, BNT162b2, and Ad26.COV2.S vaccines may be, at least in part, due to the variable immunogenicity of the vaccines described here. These data may be important in rationalizing use of the most immunogenic vaccines in high-risk individuals, consideration of prioritizing booster doses for those with the weakest responses to ameliorate hospitalization and death and tailoring booster plans by vaccine and prior infection.

Among individuals with prior infection, mRNA vaccination conferred higher antibody concentration titers. This supports recent data suggesting a single dose of mRNA vaccine in seropositive convalescent patients elicits comparable antibody titers to seronegative individuals who receive two doses of mRNA vaccine[19]. However we extend these findings by showing that this trend applies to Ad26.COV2.S and is confirmed for all three vaccines with neutralization titers regardless of vaccine type and whether a second dose was given. These findings may explain why vaccination in the setting of prior infection appears to be associated with enhanced protection[20]. In settings with limited vaccine supply, consideration to prioritizing individuals without prior infection may be warranted.

We found differences in the immunogenicity and effectiveness of the three FDA EUA vaccines in the USA. The higher immunogenicity and significantly enhanced effectiveness of mRNA1273 compared with BNT162b2 may be due to the roughly 3.3-fold higher dose administered in the mRNA1273 regimen. Surprisingly, we observed sustained lower immunogenicity of the Ad26.COV2.S vaccine by multiple measures. These findings contrast with small studies from the manufacturerClick or tap here to enter text. which employed a viral variant that was more readily neutralized *in vitro* than ancestral SARS CoV-2 here and in prior studies but concur with recent studies by Tada and colleagues[13]. These data further highlight the need for immunogenicity assessment using consistent assays to permit comparisons. Consistent with the lower efficacy in trials, breakthrough infection, hospitalization and death rates after Ad26.COV2.S were significantly higher than after mRNA1273. Additional ‘booster’ doses have already been recommended for Ad26.COV2.S recipients, regardless of other host factors in several settings notably in South Korea. Regardless of vaccine, *in vitro* neutralization of SARS CoV-2 beta variant was markedly impaired as has been previously noted. Variants that combine the neutralization evasion (as for the beta variant), with enhanced transmissibility and pathogenicity (as for the delta variant) may pose particular challenges to vaccine-induced immunity. Collectively, these data justify careful consideration of how to ensure similarly high levels of protection taking into account the primary vaccine series for individuals, in the setting of evolving variants.

The precise immune correlate of protection has yet to be agreed upon but growing data indicates the utility of anti-spike antibody titers or pseudoneutralization. A role for T-cells continues to be explored. The data here are consistent with the central role of CD4+ T-helper cells in antibody induction. However the marked heterogeneity and low frequency of cytotoxic T-cells in this, and other studies[21,22], makes this specific measure unlikely to be a universal mechanism of protection from severe disease which we note is seen at very high levels regardless of vaccine.

Several important limitations of this study are worth highlighting. First, the immunogenicity studies were focused largely healthy individuals. Further study in the particular risk groups may be warranted. Secondly, we did not measure other features of the immune system such as non-neutralizing antibodies[23] and were limited in being able to examine kinetics of responses without repeat measures. Thirdly, the effectiveness studies presented are observational and are not, by virtue of what data are available, reported stratified by age or adjusted for important potential confounders. In the absence of randomized trials comparing these vaccines directly, immunogenicity and observational effectiveness studies will continue to be required. Monitoring of vaccine immunogenicity and effectiveness could be an important strategy to identify individuals at high risk of breakthrough infection.

Taken together, these data demonstrate marked variation in the immunogenicity and effectiveness of the three FDA EUA vaccines deployed in the USA and may inform rational and equitable vaccination policy in the USA and elsewhere.

## Supporting information

Online supplement for meta-analysis of vaccine effectiveness

## Data Availability

Primary data is not available per the limited consent obtained from patients. Summary data are available by writing to the authors

## Acknowledgements

We thank Andrea Nixon and the MGH Core laboratory for assistance in performing assays. We wish to thank Michael Farzan, PhD for providing ACE2-expressing 293T cells and Aaron Schmidt for providing proteins for development of ELISA assays and Atul Bhan, MD and Mandakolathur Murali, MD for useful and insightful discussion. The RBD ELISA assay was developed by Steve Mullenbrock, Diego Farfan-Arribas, Mark Stump, Nels Pederson, Randy Wetzel and Roberto Polakiewicz of Cell Signaling Technologies, Danvers, MA. We also thank Kellie Burke, Susan Gonzalez, Dominic Iafrate, Chioma Agugoesi, Katie Tarbox, Kirsten Dickens, Pedro Ojeda, JeanCarlos Cruz, Julian Villalba, Michelle Garlin, Daniel Montes, Karla Gonzalez, Hugh Shirley, Flor Amaya, Mimi Graney, Daniel Cortez and Minjee Kim for assisting in participant recruitment. D.J.G., M.C.P., and M.N.P. were supported by the VIC Innovation fund. A.J.I. and this study were supported by the Lambertus Family Foundation. G.D.G. is supported by a Burroughs Wellcome Career Award in Medical Sciences. A.B.B. was supported by the National Institutes for Drug Abuse (NIDA) Avenir New Innovator Award DP2DA040254, the MGH Transformative Scholars Program as well as funding from the Charles H. Hood Foundation.

