## Supplementary figures and tables for "Comparative immunogenicity and effectiveness of mRNA-1273, BNT162b2 and Ad26.COV2.S COVID-19 vaccines"

**Figure S1: Immunogenicity of SARS CoV-2 vaccines by enzyme-linked immunosorbent assay (ELISA)** (described and validated in Garcia-Beltran et al. ^1^) of anti-receptor binding domain (RBD) antibodies. Data are shown according to secondary detection isotype specificity. Panel A shows IgG/M/A total antibody, Panel B shows IgG, Panel C shows IgM and panel D shows IgA. In each graph the horizontal line denotes the geometric mean concentration, and whiskers extend to 95% confidence interval. The cutoff was empirically determined based on unvaccinated individuals. ^1^) of anti-receptor binding domain (RBD) antibodies. Data are shown according to secondary detection isotype specificity. Panel A shows IgG/M/A total antibody, Panel B shows IgG, Panel C shows IgM and panel D shows IgA. In each graph the horizontal line denotes the geometric mean concentration, and whiskers extend to 95% confidence interval. The cutoff for each was empirically determined based on unvaccinated individuals.

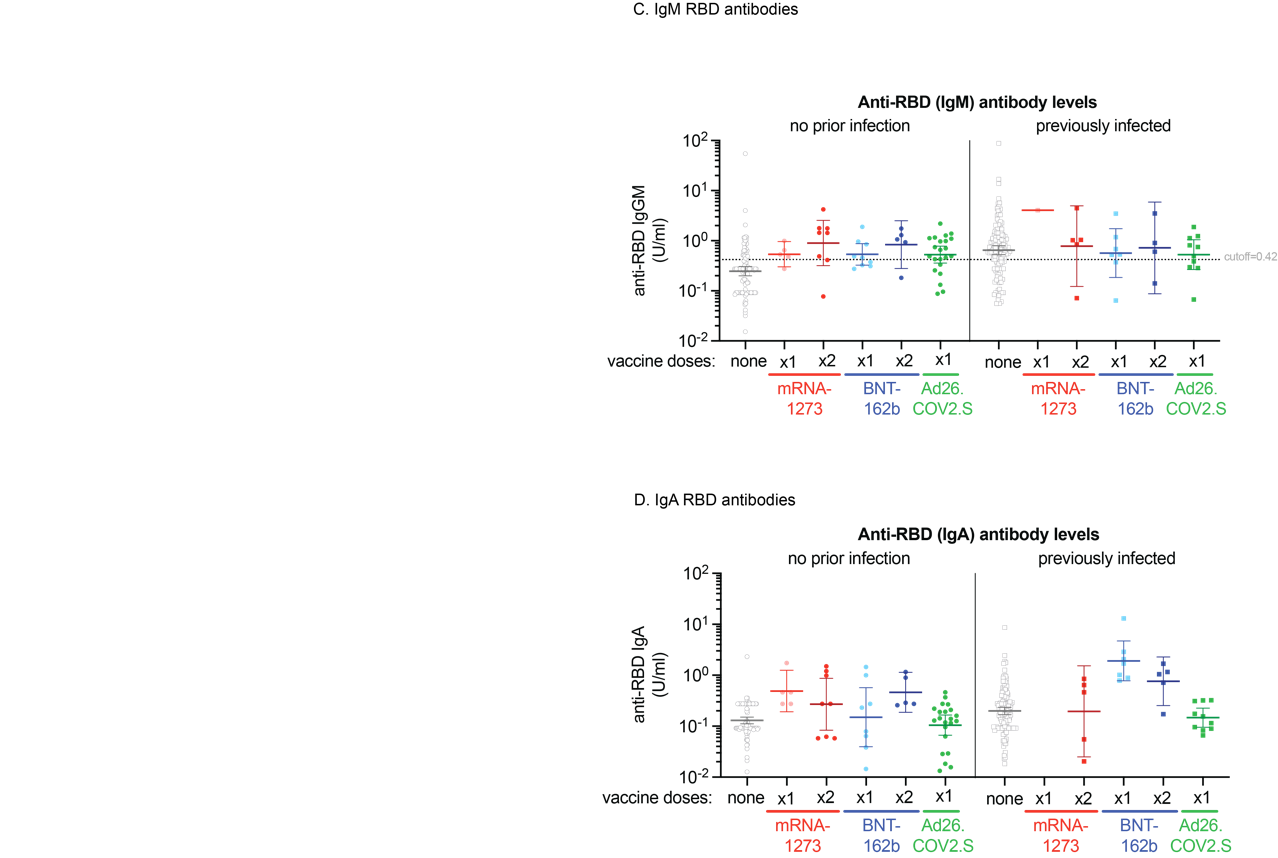

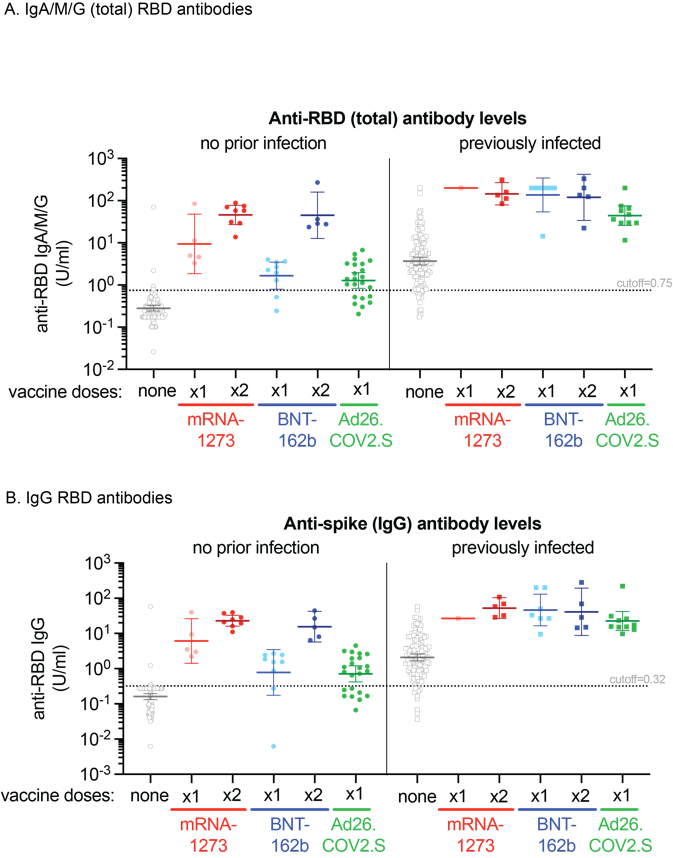

**Figure S2: Immunogenicity of SARS CoV-2 vaccines against SARS CoV-2 receptor binding domain (RBD) pentamer.** The horizontal line denotes the geometric mean concentration, and whiskers extend to 95% confidence interval.

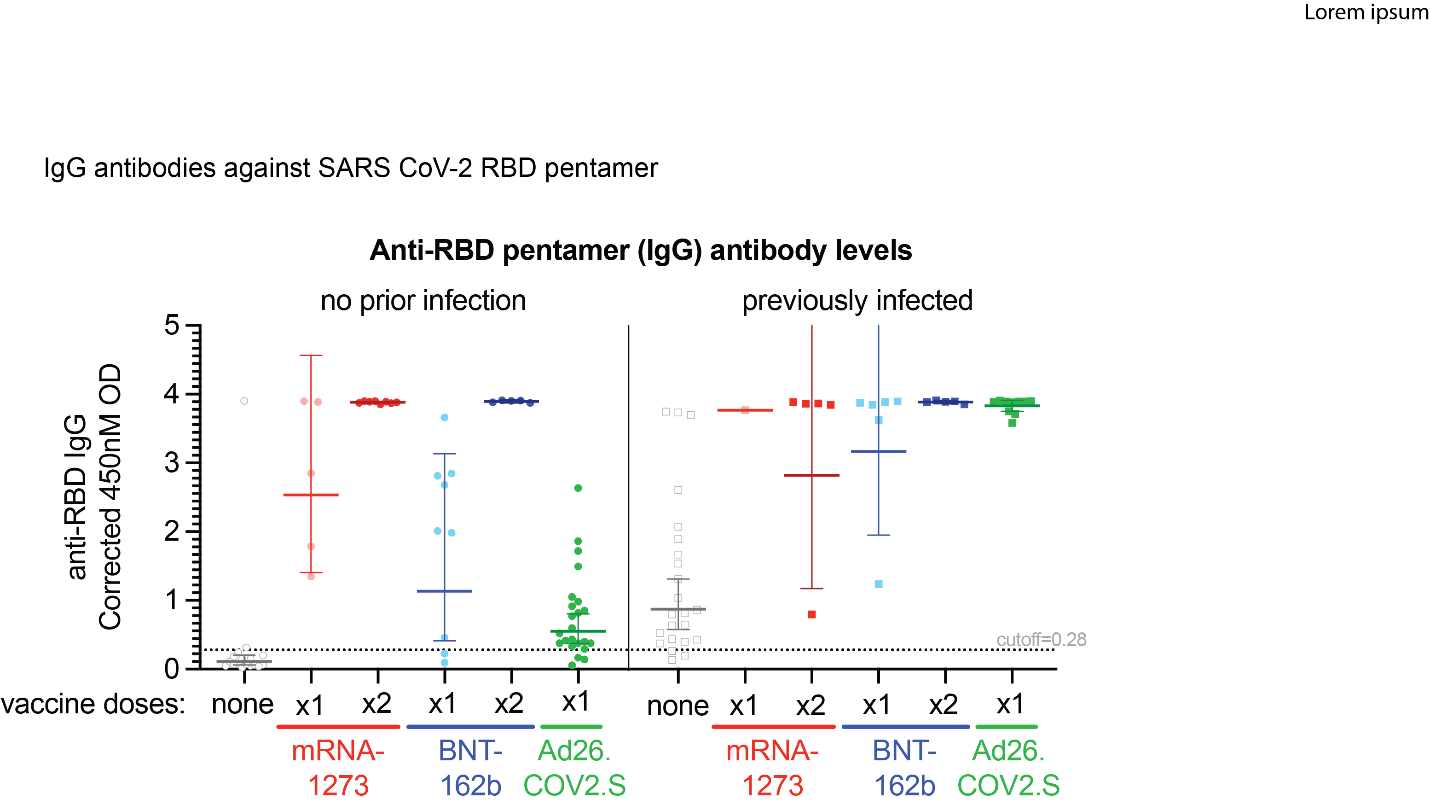

**Figure S3: Antibodies directed against RBD multimers from OC43 and HKU1 coronaviruses.** The horizontal line denotes the geometric mean concentration, and whiskers extend to 95% confidence interval.

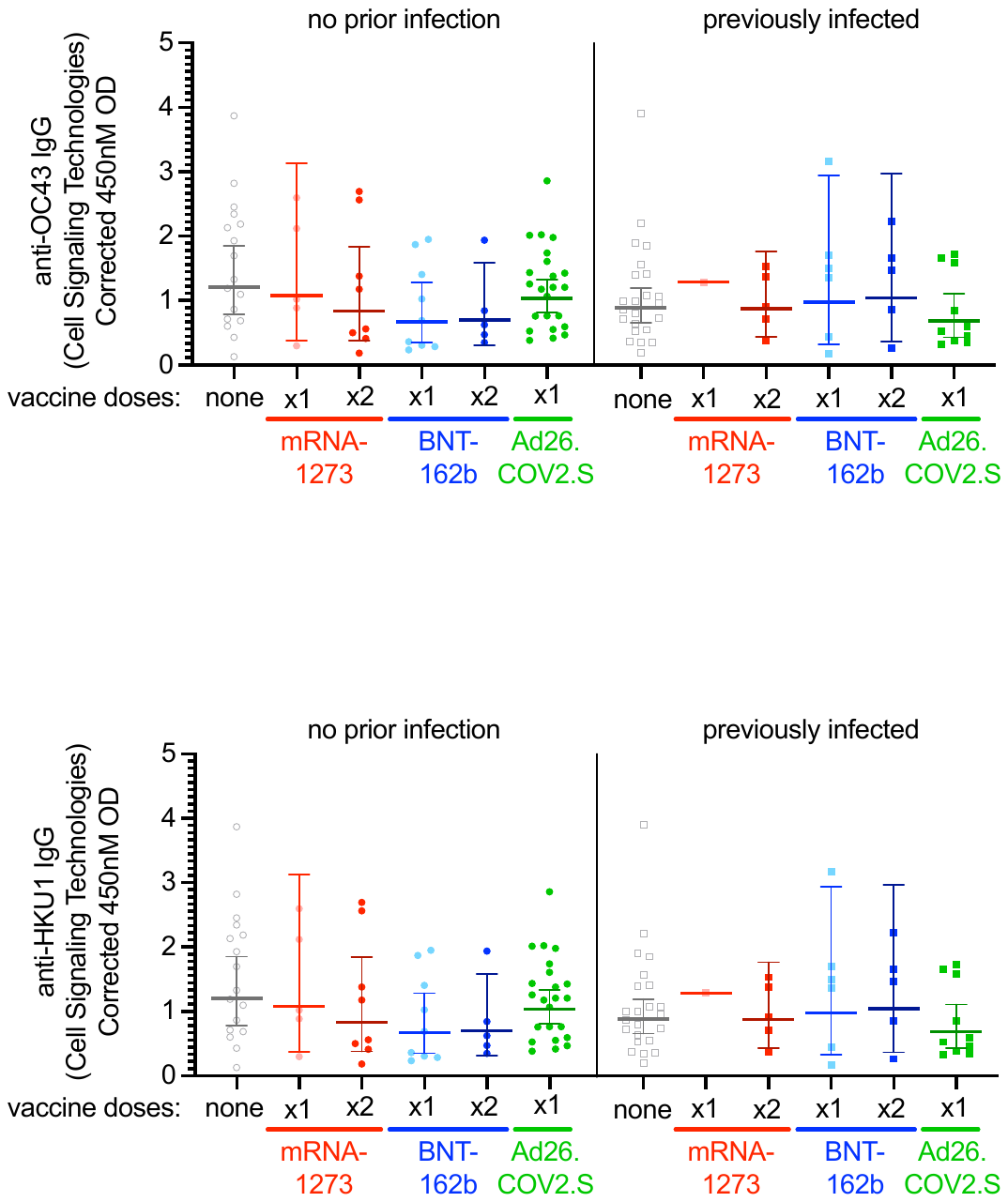

**Figure S4:** **Relative neutralization of SARS CoV-2 vaccines against SARS CoV-2 Wuhan (WT) and Victoria strains.** Analysis of serum from mRNA-1273, BNT162b2 and Ad26.COV2.S vaccinees in pseudotype neutralization assay, using the Wuhan (WT) SARS CoV-2 strain spike protein and the Victoria strain spike protein. Samples from the same subject are connected by a straight line. Fold change in geometric mean and p-values are shown.

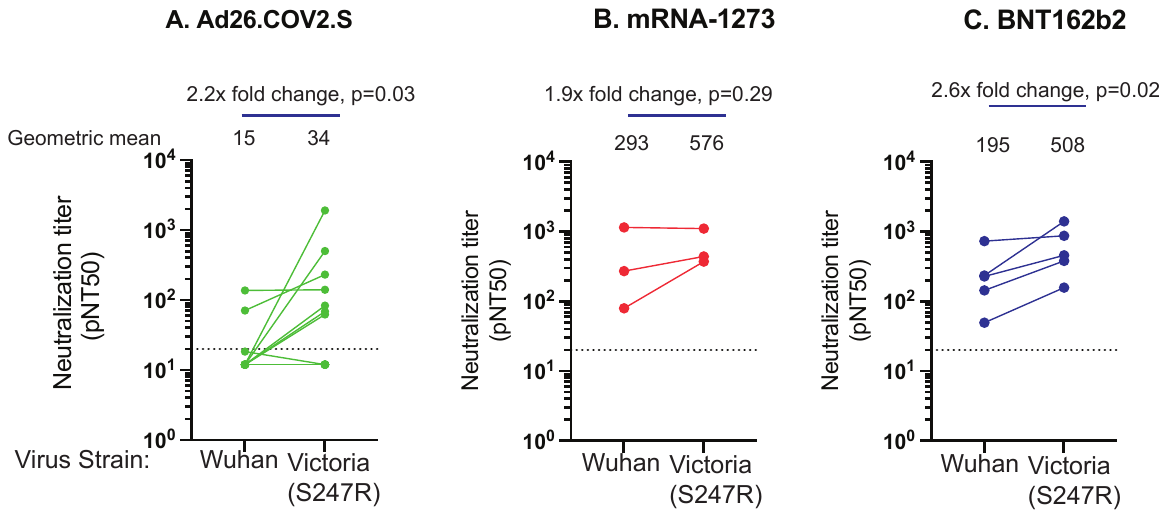

### Supplementary Tables

#### Table S1: Age, sex and duration after vaccination at time of sampling, for participants included in this study.

|  |  | **No Prior infection** | | | | | | | **Prior SARS CoV-2 infection** | | | | | |
| --- | --- | --- | --- | --- | --- | --- | --- | --- | --- | --- | --- | --- | --- | --- |
|  | **All groups** | **pre-pandemic controls** | **No vaccine** | **mRNA-1273, one dose** | **mRNA-1273, two doses** | **BNT162b2, one dose** | **BNT162b2, two doses** | **Ad26.COV2.S, one dose** | **No vaccine** | **mRNA-1273, one dose** | **mRNA-1273, two doses** | **BNT162b2, one dose** | **BNT162b2, two doses** | **Ad26.COV2.S, one dose** |
| **n** | 1,677 | 1220 | 112 | 30 | 55 | 21 | 46 | 22 | 130 | 5 | 7 | 7 | 8 | 14 |
| **Age*** |  | 36 (30-52) | 37 (31-54) | 36 (28-53) | 51 (31-60) | 32 (28-52) | 34 (31-42) | 43 (32-54) | 45 (34-53) | 35 (32-38) | 44 (36-50) | 52 (43-60) | 48 (39-58) | 42 (37-50) |
| **Sex (%)** |  |  |  |  |  |  |  |  |  |  |  |  |  |  |
| **F** | 1,092(65%) | 850 (70%) | 60 (54%) | 17 (57%) | 31 (56%) | 13 (62%) | 29 (63%) | 12 (55%) | 62 (48%) | 2 (40%) | 5 (71%) | 1 (14%) | 3 (38%) | 7 (50%) |
| **M** | 585 (35%) | 360 (30%) | 52 (46%) | 13 (43%) | 24 (44%) | 8 (38%) | 17 (37%) | 10 (45%) | 68 (52%) | 3 (60%) | 2 (29%) | 6 (86%) | 5 (62%) | 7 (50%) |
| **Days after 1st dose*** | 32 (21-48) |  |  | 24 (18-32) | 49 (46-68) | 22 (17-25) | 36 (31-53) | 24 (21-32) |  | 13 (12-20) | 52 (44-58) | 11 (10-20) | 52 (35-67) | 21 (21-46) |
| **Days after 2nd dose*** | 15 (8-30) |  |  |  | 20 (16-40) |  | 14 (10-32) |  |  |  | 22 (15-28) |  | 34 (8-41) |  |

*Median(IQR)

Table S2: Multivariate regression model of spike antibody titers in vaccine recipients, compared with unvaccinated individuals with prior infection.

| **Characteristic** | **Estimate of effect on log_10_ IgG/A/M spike antibody concentration** | **95% Confidence Interval** | **Adjusted *p*-value** |
| --- | --- | --- | --- |
| **Days after first dose (per 7 days)** | -0.02 | -0.05, 0.02 | 0.4 |
| **Age (per year)** | -0.01 | -0.02, 0.00 | <0.001 |
| **Sex** |  |  |  |
| F | *Ref* |  |  |
| M | 0.03 | -0.16, 0.21 | 0.8 |
| **Group** |  |  |  |
| Prior infection, no vaccine | *Ref* |  |  |
| No prior infection |  |  |  |
| **mRNA-1273, one dose** | -0.2 | -0.71, 0.30 | 0.4 |
| **mRNA-1273, two doses** | 1.4 | 0.88, 2.0 | <0.001 |
| **BNT162b2, one dose** | -0.7 | -1.2, -0.17 | 0.01 |
| **BNT162b2, two doses** | 0.87 | 0.35, 1.4 | 0.001 |
| **Ad26.COV2.S, one dose** | -1.7 | -2.2, -1.1 | <0.001 |
| Prior infection |  |  |  |
| **mRNA-1273, one dose** | 1.9 | 1.2, 2.6 | <0.001 |
| **mRNA-1273, two doses** | 2.1 | 1.4, 2.8 | <0.001 |
| **BNT162b2, one dose** | 1.9 | 1.2, 2.6 | <0.001 |
| **BNT162b2, two doses** | 1.7 | 1.1, 2.4 | <0.001 |
| **Ad26.COV2.S, one dose** | 1.4 | 0.86, 2.0 | <0.001 |

#### Table S3: Multivariate regression model of spike antibody titers amongst recipients of a single dose of mRNA-1273, BNT172b or Ad26.COV2.S (reference).

| **Characteristic** | **Estimate of effect on log^10^ IgG/A/M spike antibody concentration** | **95% Confidence Interval** | **Adjusted *p*-value** |
| --- | --- | --- | --- |
| **Days after first dose (per day)** | 0.07 | 0.05, 0.09 | <0.001 |
| **Age (per year)** | -0.02 | -0.03, 0.00 | 0.006 |
| **Sex** |  |  |  |
| F | *Ref* |  |  |
| M | 0.14 | -0.20, 0.48 | 0.4 |
| **Group** |  |  |  |
| No prior infection |  |  |  |
| **Ad26.COV2.S, one dose** | *Ref* |  |  |
| **mRNA-1273, one dose** | 1.5 | 1.1, 1.9 | <0.001 |
| **BNT162b2, one dose** | 1.3 | 0.88, 1.8 | <0.001 |

#### Table S4: Multivariate regression model of virus neutralization

| **Characteristic** | **Estimate of effect on log^10^ pseudovirus neutralization titre 50 (pNT50)** | **95% Confidence Interval** | **Adjusted *p*-value** |
| --- | --- | --- | --- |
| **Days after first dose (per 7 days)** | 0 | 0.00, 0.00 | 0.033 |
| **Age (per year)** | -0.01 | -0.01, 0.00 | 0.001 |
| **Sex** |  |  |  |
| F | — | — |  |
| M | -0.01 | -0.16, 0.14 | 0.9 |
| **Group** |  |  |  |
| Prior infection, no vaccine | *Ref* |  |  |
| No prior infection |  |  |  |
| **mRNA-1273, one dose** | 0.3 | -0.24, 0.83 | 0.3 |
| **mRNA-1273, two doses** | 1.3 | 0.74, 1.9 | <0.001 |
| **BNT162b2, one dose** | 0.19 | -0.35, 0.74 | 0.5 |
| **BNT162b2, two doses** | 1.3 | 0.72, 1.8 | <0.001 |
| **Ad26.COV2.S, one dose** | -0.56 | -1.1, -0.02 | 0.043 |
| Prior infection |  |  |  |
| **mRNA-1273, one dose** | 1.8 | 1.2, 2.5 | <0.001 |
| **mRNA-1273, two doses** | 2 | 1.4, 2.7 | <0.001 |
| **BNT162b2, one dose** | 1.8 | 1.2, 2.5 | <0.001 |
| **BNT162b2, two doses** | 1.9 | 1.3, 2.5 | <0.001 |
| **Ad26.COV2.S, one dose** | 1.6 | 1.0, 2.2 | <0.001 |
